# Translation, Cross-Cultural Adaptation and Measurement Properties of Three Implementation Measures FIM-AIM-IAM into Bangla

**DOI:** 10.64898/2026.01.23.26344737

**Authors:** Anika Tasneem Chowdhury, Zeeba Zahra Sultana, Faika Anjum, Syed Sharaf Ahmed Chowdhury, Md Hasan Thakur, Mohammad Hayatun Nabi, Azaz Bin Sharif

## Abstract

**Introduction:** The valid measures of implementation outcomes like acceptability, appropriateness, and feasibility are vital for bridging the evidence-practice gap in healthcare. However, instruments developed in one cultural context, such as the widely used AIM, IAM, and FIM, require rigorous validation before use in new settings. This study aimed to translate, culturally adapt, and validate the Bangla versions of the AIM, IAM, and FIM for use in Bangladesh.

**Methods:** A cross-sectional study was conducted among 549 pregnant women in Bangladesh aged 18-45 years from October and November 2025 at Rajbari District Hospital, Rajbari, Dhaka, Bangladesh. Data were collected through a pre-tested semi-structured questionnaire that included socio-demographic information, and the FIM-AIM-IAM scale. Statistical analysis, including descriptive statistics and confirmatory factor analysis, was performed using R statistical software version 4.4.0.

**Result:** Item-level analysis showed that the measurement items were normally distributed which suggested measurement of the same construct. The Bangla version of the FIM-AIM-IAM demonstrated excellent internal consistency (AIM: α = 0.78, IAM: α = 0.81, FIM: α = 0.77). This version demonstrated strong construct validity, as confirmed by a confirmatory factor analysis that supports a unidimensional model with an excellent fit (CFI = 0.92, RMSEA = 0.04). The scales also demonstrated adequate convergent validity, with Average Variance Extracted (AVE) values of 0.51, 0.52, and 0.5 for FIM, AIM, and IAM, respectively.

**Conclusion:** The Bangla version of the FIM-AIM-IAM was found to be a valid screening tool for facilitating high-quality implementation research in maternal and child health settings, ultimately supporting the evidence-based adoption and sustained implementation of interventions in Bangla-speaking settings.

## INTRODUCTION

The constant gap between evidence-based research and its routine application in healthcare remains a major challenge across health systems worldwide (1,2). Implementation science has appeared as a discipline that is dedicated to addressing this gap by examining the methods, strategies, and contextual factors that facilitate or inhibit the adaptation of evidence-based practices (EBPs) into real-world care (3,4). This discipline employs a variety of theories, models, and frameworks to guide implementation efforts (5,6). A foundation of this scientific inquiry is the careful assessment of implementation success through clearly defined measures (7,8). A well-recognized taxonomy recognizes eight implementation outcomes—acceptability, appropriateness, feasibility, adoption, fidelity, implementation cost, coverage, and sustainability. Each of these represents a vital dimension of how interventions are perceived, delivered, and maintained in practice (9). Among these, acceptability, appropriateness, and feasibility are often considered early “leading indicators” that direct the probability of successful implementation (9,10). Acceptability reflects stakeholders’ approval or satisfaction with an intervention; appropriateness demonstrates perceived relevance within a specific setting or problem, and feasibility examines the extent to which an intervention can be effectively carried out as planned in a given context (9).

To optimize the implementation process, it is essential to validate and use psychometrically sound measures to accurately assess outcomes (7,11). Reliable data provided by such tools is vital for assessing progress, identifying barriers, and informing strategic improvements throughout the implementation process (12). To operationalize the key outcomes of implementation, three brief, four-item instruments were developed: the Acceptability of Intervention Measure (AIM), the Intervention Appropriateness Measure (IAM), and the Feasibility of Intervention Measure (FIM) (13). These measures were designed to provide pragmatic, valid data on stakeholder perception and have exhibited strong reliability and validity in their original development and testing (13). The observations from these tools are crucial because early indication of an intervention’s acceptability, appropriateness, and feasibility is believed to be important for its later adoption and long-term sustainability (9).

A foundational principle of cross-cultural research is that instruments developed in one language and cultural context cannot be presumed valid in another without vigilant adaptation to confirm conceptual, linguistic, and technical relevance (14,15). This concept directly applies to the AIM, IAM, and FIM, which were primarily developed and validated in a high-income, English-speaking context (the United States) (13). Without rigorous cross-cultural adaptation, the direct application of such instruments in linguistically and culturally diverse settings might result in conceptual non-equivalence and measurement bias, which can lead to unreliable findings (16,17). Such methodological limitations can misguide implementation efforts and contribute to a persistent equity gap in implementation science for low- and middle-income countries (LMICs) (18). The feasibility and value of cross-cultural adaptation are well-informed by international validations of these tools; German, Malay, and Brazilian-Portuguese versions have shown robust psychometric properties, including internal consistency and coherent factor structure, and also reported context-specific measures, such as the ceiling effect (19–21). These findings underscore the need for stringent adaptation to ensure the reliability, validity, and comparability of implementation outcomes across diverse settings. Despite successful adaptations in other languages,, no validated Bangla versions of the AIM, IAM, or FIM exist at present, which limits the assessment capacity of early implementation measures in Bangla-speaking populations appropriately. Given the linguistic, cultural, and healthcare system differences, the direct use of English instruments could compromise conceptual equivalence and measurement validity. Maternal health settings represent a key context for implementation research in LMICs such as Bangladesh, as pregnant women are primary recipients of multiple evidence-based interventions, and their perceptions of acceptability, appropriateness and feasibility can influence service uptake and continuity of care (22,23). To address this gap, the present study aims to translate, culturally adapt, and validate the Bangla versions of AIM, IAM, and FIM among pregnant women attending antenatal care at a district hospital in Bangladesh. This will provide psychometrically sound and contextually relevant instruments, helping to facilitate implementation research in maternal and child health settings. Finally, this will support the practical adoption and persistence implementation of interventions in Bangla-speaking settings.

## METHODOLOGY

### Study design and setting

This methodological study followed the Consensus-based Standards for the Selection of Health Measurement Instruments (COSMIN) framework (14). It involved cross-cultural adaptation and psychometric evaluation of three implementation outcome measures developed by (13): the Acceptability of Intervention Measure (AIM), Intervention Appropriateness Measure (IAM), and Feasibility of Intervention Measure (FIM) (2). Each scale comprises four items rated on a 5-point Likert scale (1 = completely disagree to 5 = completely agree), yielding total scores from 4 to 20, with higher scores indicating greater acceptability, appropriateness, or feasibility. Data were collected between October and November 2025 at Rajbari District Hospital, Rajbari, Dhaka, Bangladesh, among pregnant women attending antenatal care. We have followed the STROBE guideline to report this study [**Supplementary Material 1**].

This study was implemented in three stages: (1) translation and cross-cultural adaptation of the AIM, IAM, and FIM into Bangla; (2) field administration of the adapted instruments; and (3) evaluation of measurement properties.

### Eligibility criteria

Eligible participants were pregnant women aged 15–49 years who attended antenatal care and could read and communicate in Bangla. Women with cognitive impairment or severe mental health conditions compromising consent or questionnaire completion were excluded, as were those who were critically ill, had high-risk pregnancies requiring urgent intervention, declined participation, or, in the case of minors, lacked guardian consent and/or assent.

### Stage 1: Translation and cross-cultural adaptation

Translation and adaptation were conducted in accordance with internationally accepted guidelines for patient-reported outcome measures. Six native Bangla-speaking translators independently translated the AIM, IAM, and FIM into Bangla, and two bilingual translators, blinded to the study objectives, independently back-translated the preliminary Bangla versions into English. Back-translations were compared with the originals to verify equivalence.

An expert panel of ten individuals (one measurement specialist, five public health professionals, four subject-matter experts) evaluated the translated items using a 5-point rating scale for relevance, clarity, and consistency with the intended construct. Face validity was assessed using a convenience sample of 20 pregnant women, who rated the items for readability and comprehensibility on a 5-point Likert scale (1 = not relevant, 5 = highly relevant). Based on expert ratings, participant feedback, and comparison of original and back-translated versions, the final Bangla versions of the AIM, IAM, and FIM were produced. The final translated version is provided as **Supplementary Material 2**.

### Stage 2: Data collection procedures

In field testing, the finalized Bangla AIM, IAM, and FIM were self-administered by 549 pregnant women after antenatal consultations. Data collectors provided standardized instructions and assisted with procedural queries. For the test–retest reliability, a simple random subsample of about 10% (n = 50) completed the three scales again after a 7-day interval. Additional socio-demographic data, including age, education, and occupation, were collected using a structured questionnaire.

### Stage 3: Assessment of measurement properties

Data was entered into Microsoft Excel and analyzed using *R statistical software version 4.4.0*. Frequency (n) and percentage (%) were used to summarize the categorical data for descriptive analysis. Means, standard deviations, skewness, kurtosis, item-total correlations, and Cronbach’s alpha of the scale if each item was deleted were calculated and reported. Interpretability was examined by analyzing score distributions and floor and ceiling effects, defined as present if at least 15% of participants obtained the minimum or maximum score. The proportion of missing responses for each item was calculated as an indicator of the acceptability of the survey. Internal consistency of each scale was assessed using Cronbach’s alpha (acceptable range 0.70–0.95). Test–retest reliability in the subsample was evaluated using the intraclass correlation coefficient (ICC). Measurement error was quantified using the Standard Error of Measurement (SEM) and the Smallest Detectable Change (SDC). Construct validity was examined with the Confirmatory Factor Analysis (CFA) to test the hypothesized unidimensional structure of each scale. Model fit was evaluated using the Comparative Fit Index (CFI), Tucker-Lewis Index (TLI), and Root Mean Square Error of Approximation (RMSEA). Convergent validity was assessed by calculating the Average Variance Extracted (AVE).

### Sample size

The sample size followed the recommendations for factor-analytic and psychometric studies. Hair et al. suggest a minimum of 100 participants (3), and Comrey and Lee classify 500 as a good sample (13). The target sample was therefore set at minimum 500.

### Quality assurance

Data collectors received training to standardize administration and recording. Completed questionnaires were reviewed, and missing or unclear responses were clarified when possible. Data were double-entered, discrepancies were resolved by cross-checking, and the final dataset underwent checks before analysis.

### Ethical considerations

Ethical approval was obtained from the Institutional Review Board/Ethical Review Committee of the Department of Public Health, North South University, Bangladesh (reference: 2025/OR-NSU/IRB/1011). Written informed consent was obtained from all participants; for minors, consent from a guardian and assent from the participant were required. Participation was voluntary, with the option to withdraw at any time without consequences for care. Personal identifiers were removed or coded to ensure confidentiality, and the study adhered to the principles of the Declaration of Helsinki.

## RESULTS

### General characteristics of the participants

**Table 1** presents the descriptive characteristics of the 549 study participants. The cohort had a mean age of 23.39 years (±5.19), and the vast majority (98.29%) were not working. Most participants (83.06%) and their husbands (63.60%) had attained a secondary or higher secondary education level.

**Table 1:** Descriptive characteristics of the study participants.

| **Table 1:** Descriptive characteristics of the study participants |  |  |
| --- | --- | --- |
| **Variable** | **Categories** | **Mean (±SD) / N (%)** |
| Age | | 23.39 (±5.19) |
| Education level of the participants | Primary/No Education | 60 (10.93) |
|  | Secondary/Higher Secondary | 456 (83.06) |
|  | Undergraduate | 10 (1.82) |
|  | Graduate | 23 (4.19) |
| Husband's education level | Primary/No Education | 131 (24.08) |
|  | Secondary/Higher Secondary | 346 (63.60) |
|  | Undergraduate | 32 (5.88) |
|  | Graduate | 35 (6.43) |
| Working status of the participant | Not working | 540 (98.29) |
|  | Working | 9 (1.64) |

### Item-level analysis

**Table 2** presents item-level statistics for the Bangla FIM, AIM, and IAM. All items of the three domains exhibited high mean scores from 4.14 to 4.39 on a 5-point Likert scale. This indicates a strong positive perception of the intervention. Little variability was observed in responses, as the standard deviations (SD) were low (0.35 to 0.53). The item-total correlations were strong (>0.60) for all components, indicating that each item correlates well with its respective scale’s total score. The Cronbach’s alpha value for each scale remained high (range: 0.890 to 0.898) even after an item was deleted, indicating that all items contribute meaningfully to their respective constructs. The majority of the skewness (−0.13 to 1.92) and kurtosis (−0.04 to 2.15) values for each item are within an acceptable range.

**Table 2:** Mean, standard deviation, item-total correlation, skewness, kurtosis, and Cronbach’s alpha of the scale for each translated scale item.

|  |  |  |  |  |  |  |
| --- | --- | --- | --- | --- | --- | --- |
| Item | Mean | SD | Skewness | Kurtosis | Item total correlation | Alpha If deleted |
| **Acceptability** |  |  |  |  |  |  |
| Item1 | 4.14 | 0.35 | 1.92 | 2.15 | 0.64 | 0.894 |
| Item2 | 4.17 | 0.38 | 1.64 | 1.03 | 0.70 | 0.892 |
| Item3 | 4.15 | 0.36 | 1.86 | 1.90 | 0.65 | 0.894 |
| Item4 | 4.35 | 0.50 | 0.15 | -0.04 | 0.63 | 0.893 |
| **Appropriateness** |  |  |  |  |  |  |
| Item1 | 4.37 | 0.53 | -0.13 | 0.44 | 0.68 | 0.890 |
| Item2 | 4.33 | 0.48 | 0.58 | -1.27 | 0.73 | 0.898 |
| Item3 | 4.35 | 0.51 | 0.08 | -0.03 | 0.69 | 0.890 |
| Item4 | 4.31 | 0.46 | 0.84 | -1.30 | 0.64 | 0.892 |
| **Feasibility** |  |  |  |  |  |  |
| Item1 | 4.25 | 0.45 | 0.75 | 0.33 | 0.63 | 0.893 |
| Item2 | 4.26 | 0.47 | 0.50 | 0.87 | 0.61 | 0.894 |
| Item3 | 4.25 | 0.45 | 0.78 | 0.27 | 0.67 | 0.891 |
| Item4 | 4.39 | 0.51 | 0.13 | -0.79 | 0.67 | 0.891 |

### Interpretability

The ceiling and floor effects were less than 1% of participants scoring the minimum across all scales. There were no missing responses for any of the items in the finalized scales. This indicates high acceptability and comprehensibility among the participants.

### Reliability

The reliability metrics for the Bangla FIM, AIM, and IAM are summarized in **Table 3**. Cronbach’s alpha was used to measure internal consistency, which remained adequate for all three scales (AIM: α = 0.78, IAM: α = 0.81, FIM: α = 0.77). The test-retest reliability, assessed over a 7-day interval in a subsample of 50 participants, was moderate. The intraclass correlation coefficients (ICCs) were calculated as 0.41 (95% CI: 0.14–0.62) for AIM, 0.52 (95% CI: 0.28–0.69) for IAM, and 0.49 (95% CI: 0.25–0.68) for FIM. The Standard Error of Measurement (SEM) was low for all sub-scales that varied from 0.88 (5.55% of total score) for AIM to 1.13 (7.08% of total score) for FIM. The corresponding Smallest Detectable Change (SDC) values were 2.46 for AIM, 3.05 for IAM, and 3.14 for FIM. The composite reliability was estimated to be 0.79, 0.81, 0.77 for AIM, IAM, and FIM, which is above the threshold of 0.7.

**Table 3:** Reliability of three implementation outcome measures (AIM, IAM, and FIM)

|  |  |  |  |  |  |
| --- | --- | --- | --- | --- | --- |
| **Domain** | **Cronbach Alpha** | **ICC (95% CI)** | **SEM, % related to total score** | **SDC** | **CR** |
| AIM | 0.78 | 0.41 (0.14 – 0.62) | 0.88, 5.55 | 2.46 | 0.79 |
| IAM | 0.81 | 0.52 (0.28 – 0.69) | 1.10, 6.88 | 3.05 | 0.81 |
| FIM | 0.77 | 0.49 (0.25 – 0.68) | 1.13, 7.08 | 3.14 | 0.77 |

### Validity

Face validity was assessed and confirmed via an expert review process involving both subject matter specialists and key stakeholders. The construct validity of the scales was assessed. An initial exploratory factor analysis (EFA) demonstrates a dominant single factor. As shown in **Table 4**, the first factor had an eigenvalue of 5.78, explaining 48% of the total variance, and the second factor had an eigenvalue of 1.22, explaining an additional 10%. The scree plot criterion also suggested a single-factor solution. Subsequent Confirmatory Factor Analysis (CFA) was performed to test this unidimensional model for the combined 12-item scale. **Figure 2** demonstrates the CFA for the combined Bangla FIM-AIM-IAM scale. It shows a single-factor model where all 12 items (qa1 to qa4, qb1 to qb4, and qc1 to qc4) load have one underlying construct. The standardized loadings ranged from 0.64 to 0.77. The high correlations between the original latent constructs (Acceptability, Appropriateness, and Feasibility). This also shows a perfect correlation (1.00) between Appropriateness and Feasibility, which provides statistical evidence that these concepts are not distinct in this sample. **Table 5** and **Figure 1** demonstrate an excellent model fit. The model met the criteria for sample adequacy (KMO = 0.92) and sphericity (Bartlett’s test, p < 0.001). The absolute fit indices were strong (χ²/df = 2.01, RMSEA = 0.04, SRMR = 0.06, GFI = 0.91). The incremental fit indices also met or were approximate to acceptable thresholds (CFI = 0.92, NFI = 0.91, TLI = 0.87, AGFI = 0.87). All factor loadings in the model were statistically significant and further support the single-factor structure. Average Variance Extracted (AVE) was also calculated to estimate the convergent validity, which was found to be 0.51 for AIM and 0.52 for IAM (0.52) scales and 0.50 for FIM.

**Figure 1:**
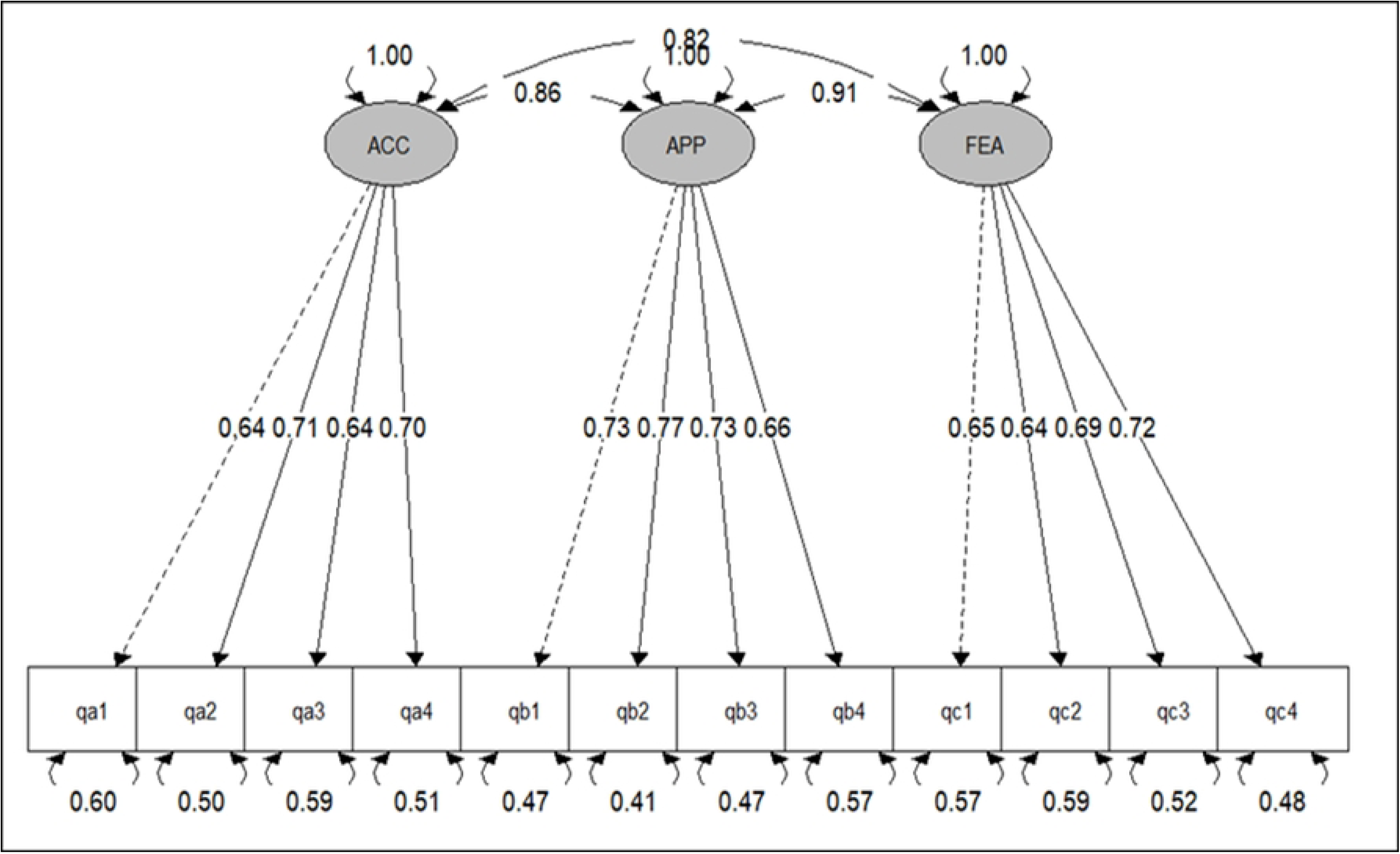
Structural equation modeling (SEM) of the FIM-AIM-IAM

**Table 4:**
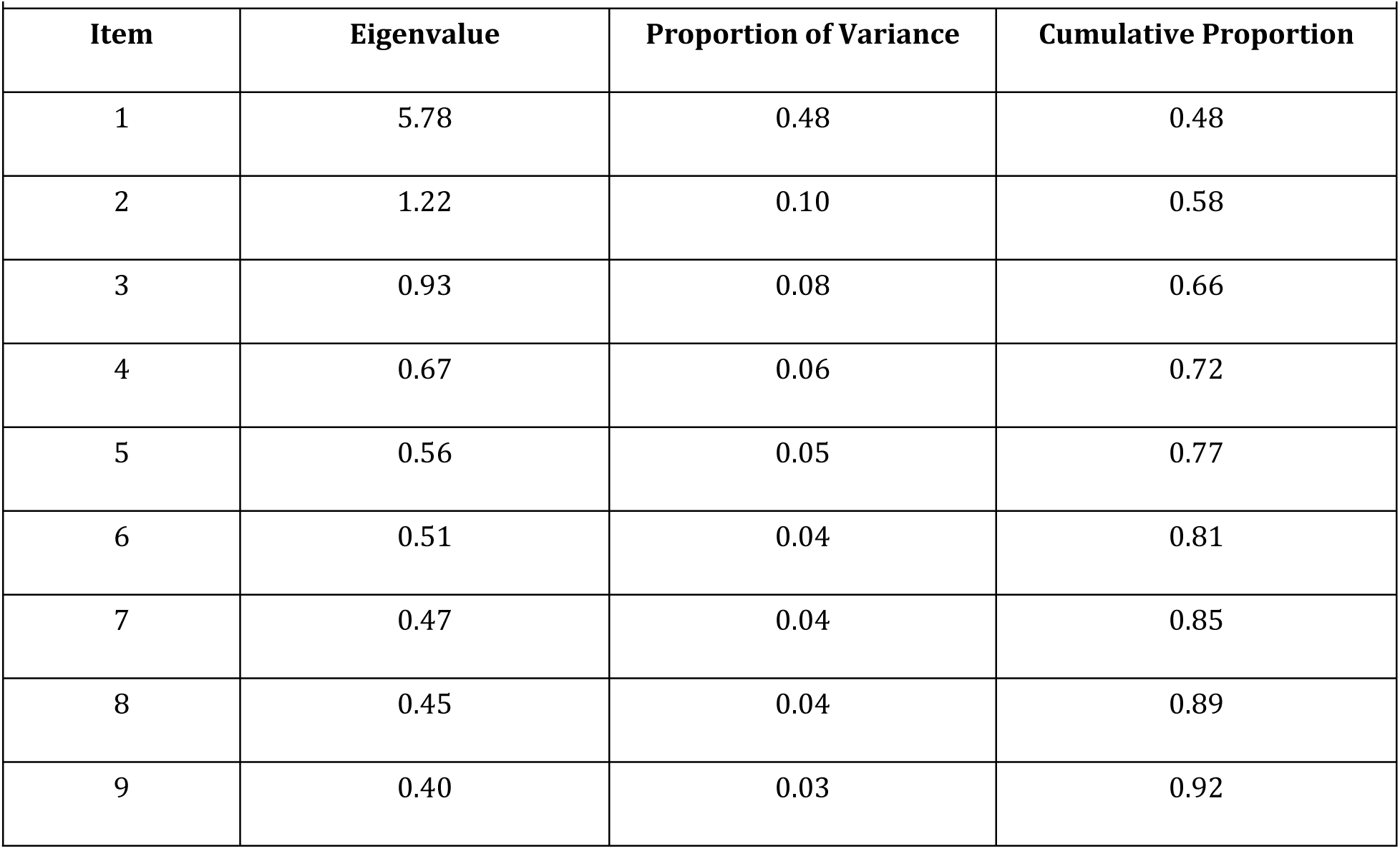
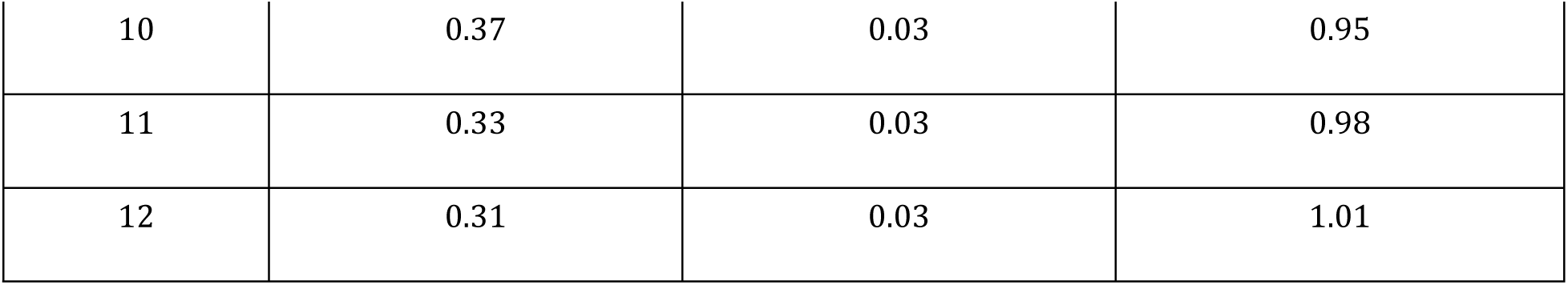
Eigenvalue and total variances explained for the final single structure.

**Table 5:**
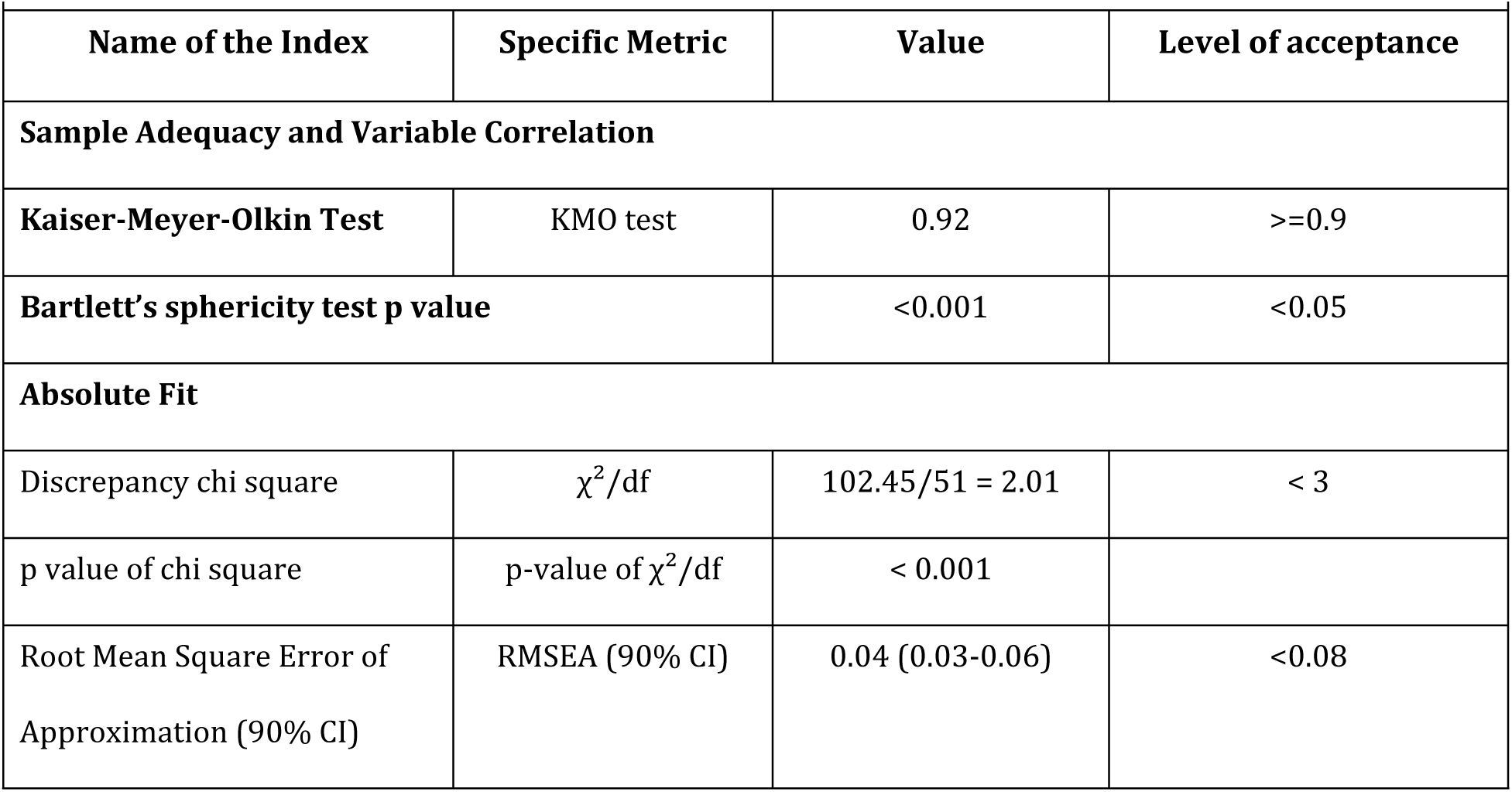
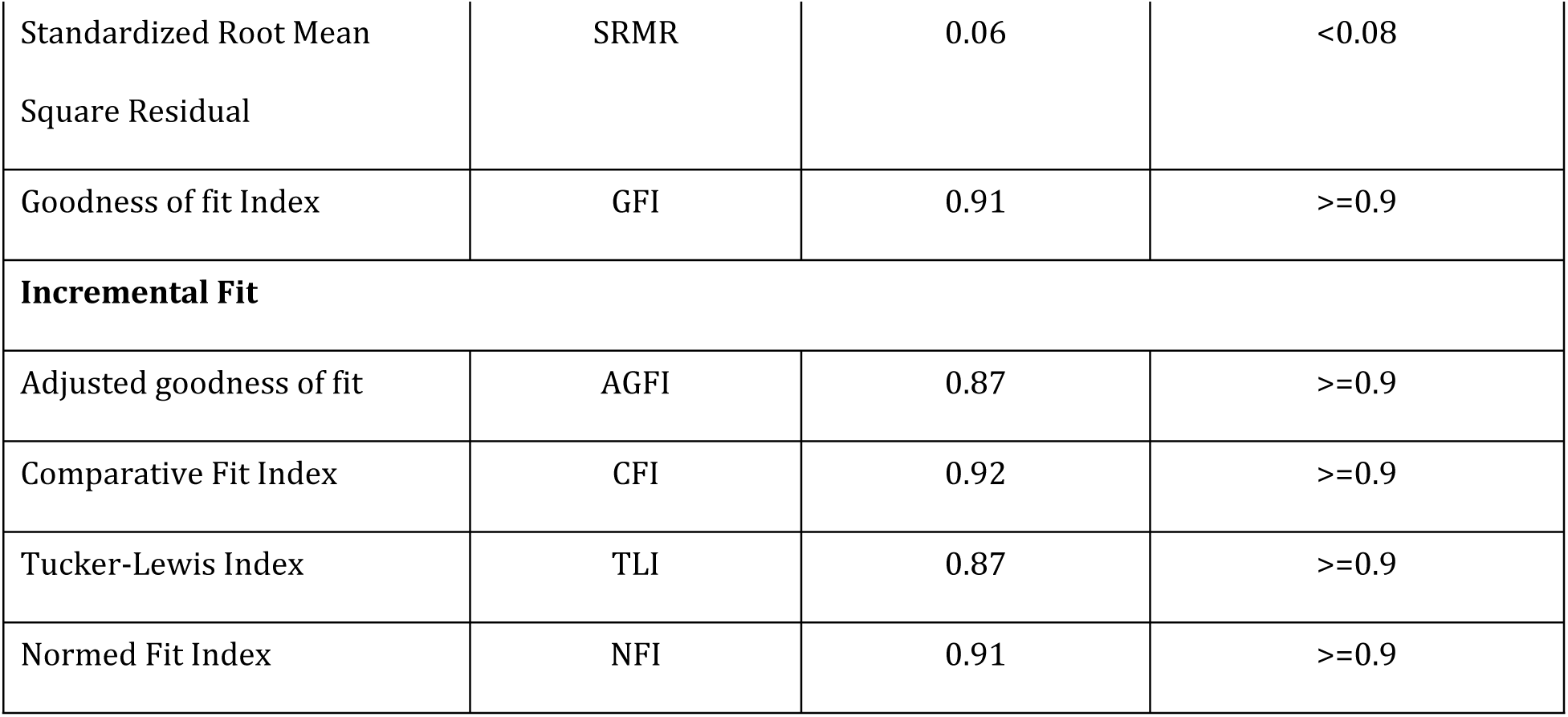
Psychometric properties of the Bangla FIM-AIM-IAM.

| Table 5: Psychometric properties of the Bangla FIM-AIM-IAM |  |  |  |
| --- | --- | --- | --- |
| Name of the Index | Specific Metric | Value | Level of acceptance |
| **Sample Adequacy and Variable Correlation** |  |  |  |
| Kaiser-Meyer-Olkin Test | KMO test | 0.92 | $\geq 0.9$ |
| Bartlett's sphericity test p value |  | <0.001 | <0.05 |
| **Absolute Fit** |  |  |  |
| Discrepancy chi square | $\chi^2/\text{df}$ | 102.45/51 = 2.01 | < 3 |
| p value of chi square | p-value of $\chi^2/\text{df}$ | < 0.001 | |
| Root Mean Square Error of Approximation (90% CI) | RMSEA (90% CI) | 0.04 (0.03-0.06) | <0.08 |
| Standardized Root Mean Square Residual | SRMR | 0.06 | <0.08 |
| Goodness of fit Index | GFI | 0.91 | >=0.9 |
| **Incremental Fit** |  |  |  |
| Adjusted goodness of fit | AGFI | 0.87 | >=0.9 |
| Comparative Fit Index | CFI | 0.92 | >=0.9 |
| Tucker-Lewis Index | TLI | 0.87 | >=0.9 |
| Normed Fit Index | NFI | 0.91 | >=0.9 |

### Scale-level psychometric properties

The psychometric properties of the Bangla FIM-AIM-IAM are presented in **Table 5**. The scale showed excellent sample adequacy for factor analysis. The combined model demonstrated a good absolute fit (RMSEA = 0.04, SRMR = 0.06, GFI = 0.91) and a satisfactory incremental fit (CFI = 0.92, NFI = 0.91). The internal consistency for the overall 12-item scale was high (Cronbach’s alpha = 0.92).

## DISCUSSION

This study aimed to translate, cross-culturally adapt, and validate the FIM, AIM, and IAM implementation outcome measures in the Bangla language. The findings demonstrate that the Bangla versions of these scales are valid and reliable tools for assessing the acceptability, appropriateness, and feasibility of health interventions in the study context. The 12-item scale showed a good model fit and acceptable reliability, which supports its potential for use in implementation research.

The translation and adaptation process was meticulous, following established guidelines (14). This resulted in a version that was easily understood by the target population, which was evidenced by the absence of any missing data. The item-level analysis confirmed that all items were well-performing and had a high mean score, which indicates a strong positive understanding of the intervention under study. The strong item-total correlations (>0.60) suggest that each item contributes significantly to its respective construct of the scale (24). The skewness and kurtosis values for most items fell within an acceptable range which suggests a relatively normal distribution.

Regarding reliability, the internal consistency of all three domains of the implementation scales was acceptable. The Cronbach’s alpha values were found to be all above the recommended threshold of 0.70 for group-level analysis (25). These values are slightly lower than those reported in the original English version (AIM: 0.85, IAM: 0.91, FIM: 0.89) by Weiner et al. (13) but are comparable to other validation studies (21). The test-retest reliability, however, was moderate with ICCs between 0.41 and 0.52 (26), which is lower than the substantial to excellent reliability found in the original study, with ICCs ranging from 0.83 to 0.88 (13). This could be due to actual changes in participants’ perceptions over the 7–14-day retest period within the dynamic context of a treatment program, or it may reflect a limitation in the stability of the measures in this specific population. The low SEM and SDC values are positive indicators that suggest the tools are precise enough to detect meaningful changes in scores.

The construct validity of the Bangla FIM-AIM-IAM was strongly supported. Contrary to the original three-factor structure (13), our EFA and CFA supported a robust single-factor model. This results concludes that a unidimensional model best fits the data, and it also indicates that Bangla-speaking participants perceive acceptability, appropriateness, and feasibility as a single concept. The model shows significant relationships between all items and the single factor. This finding was found to be in line with a previous study conducted in Malaysia (20) that indicates, in some cultural contexts, the theoretical distinctions between these implementation outcomes may not hold factual. The CFA indices (CFI=0.92, RMSEA=0.04, SRMR=0.06) demonstrated an excellent model fit, with all values exceeding the conventional threshold (27). This result provided a strong statistical support for this unidimensional structure in the study sample.

## Acknowledgements

We sincerely thank all participants who generously contributed their time to this study.

## Availability of data and materials

The de-identified dataset can be found in the source https://doi.org/10.6084/m9.figshare.30926762

